# The epidemiology of periportal fibrosis and relevance of current *Schistosoma mansoni* infection: a population-based, cross-sectional study

**DOI:** 10.1101/2023.09.15.23295612

**Authors:** Seun Anjorin, Betty Nabatte, Simon Mpooya, Benjamin Tinkitina, Christopher K. Opio, Narcis B. Kabatereine, Goylette F. Chami

**Author notes:** Correspondence Big Data Institute, University of Oxford, Oxford, United Kingdom OX3 7LF.

## Abstract

**Background:** Intestinal schistosome infections are known to cause periportal fibrosis (PPF). Yet, the epidemiology of PPF remains poorly understood, especially in settings endemic with *Schistosoma mansoni*.

**Methods:** We randomly sampled 1442 households from 38 villages in Mayuge, Buliisa, and Pakwach Districts of Uganda within the SchistoTrack Cohort to examine 2834 individuals aged 5-90 years. PPF was diagnosed using ultrasound and image patterns C-F from the Niamey Protocol. *S. mansoni* infection status/intensity was diagnosed by Kato-Katz microscopy and point-of-care circulating cathodic antigens (POC-CCA). Schistosome infection, coinfections, and comorbidities were examined as exposures for PPF. Logistic regressions were run with standard errors clustered by household.

**Findings:** PPF prevalence was 12·10% (343/2834), varying from 5·00-19·46% across districts. *S. mansoni* prevalence by Kato-Katz, and POC-CCA trace negative and positive was 43·37% (1229/2834), 40·86% (1158/2834), and 65·73% (1863/2834) respectively. Individual schistosome infection status/intensity was not correlated with the likelihood of PPF. Living in a village where adults had <5% prevalence of heavy intensity infections (400+ eggs per gram of stool) was associated with 30.2% decreased odds of PPF. The likelihood of PPF with age linearly increased from 5-25, exponentially changed from 26-45, remain unchanged from 45- 60, and steadily decreased past 60 years. History of liver diseases, human immunodeficiency virus positivity (HIV+), and ultrasound-detected chronic hepatitis/early cirrhosis-like disease were associated with >2-fold increased PPF likelihood.

**Interpretation:** Current individual schistosome infections alone are uninformative for PPF. History of HIV+ and underlying chronic hepatitis/early cirrhosis-like disease were risk factors and could be investigated for PPF surveillance and management.

**Funding:** Nuffield Department of Population Health Pump Priming Fund, Wellcome Trust Institutional Strategic Support Fund (204826/Z/16/Z), John Fell Fund, Robertson Foundation Fellowship, and UKRI EPSRC Award (EP/X021793/1).

**Research in context:** *Evidence before this study:* Morbidity due to parasitic infection is a complex interplay of current and past exposures. World Health Organization (WHO) guidelines for elimination of schistosomiasis as a public health problem assume current infection is a reliable proxy indicator of prevalent morbidity. Community infection thresholds are defined in guidelines and, when met, the assumption is there is no schistosomiasis-related morbidity. There is a lack of evidence for the association of infection with prevalent morbidity in the context of repeated treatment from routine mass drug administration. To evaluate WHO guidelines, there is a need for large-scale population-based, cross-sectional studies in endemic areas where current infection is compared with current morbidity at the same timepoint. A cross-sectional design also enables the investigation of a wide range of risk factors to assess the relative importance of current schistosome infection. Periportal fibrosis is a schistosomiasis-associated severe morbidity with clinical consequences such as portal hypertension, upper gastrointestinal tract bleeding, and ultimately premature death. Yet, little is known about the distribution of this disease; no information is available on its most basic epidemiology including age and gender-specific likelihoods. It is schistosomiasis-specific or attributable to schistosome pathology unlike more subtle conditions with complex aetiologies (e.g. anaemia). Hence, investigating periportal fibrosis serves as a first line, conservative approach to evaluating World Health Organization guidelines for elimination of schistosomiasis-related morbidity as a public health problem. A systematic literature search as part of an ongoing metanalysis was prospectively registered on PROSPERO (CRD42022333919). Databases were searched on 18^th^ May 2022 and included the Cochrane Central Register of Controlled Trials, Embase, Global Health, Global Index Medicus, and Medline. The following general terms were used: “Schistosoma” AND “fibrosis” AND “intensity” AND “infection” AND “periportal OR liver”. Studies were included that considered *Schistosoma mansoni*, *S. japonicum,* or *S. mekongi* species. Only original research articles in English were considered. No restriction on date of publication, age, gender, or region was applied. Infection was required to be diagnosed as opposed to self-reported. Periportal fibrosis was defined by the study authors.

*Added value of this study:* No population-wide or adjusted analyses were found to characterize the age-specific likelihood of periportal fibrosis. Studies focused on unadjusted associations in nonrandomly sampled populations of narrow age groups. In a single case (Weigand et al 2021) where adjusted analyses were completed (with age and gender considered), national programmatic data was used with sparsely nonrandomly sampled schoolchildren and limited frequency of the outcome of periportal fibrosis. Ultrasound data collection protocols and validation across studies were poorly reported. There was a lack of investigations on coinfections and comorbidities with only eight studies initiated (or published) from 2003 onwards after the start of mass drug administration in sub-Saharan Africa.

*Implications of all available evidence:* To our knowledge, this study is the first to characterize the epidemiology of periportal fibrosis with respect to the most common intestinal schistosome pathogen (*S. mansoni*). We conducted a comprehensive, population-based study of all ages (5+ years) eligible for mass drug administration in an area that has received at least 13 rounds of treatment. Here we provide clear evidence for the lack of association of current *S. mansoni* infection status and intensity with periportal fibrosis irrespective of the diagnostic for schistosome infection. No support was found for current WHO elimination guidelines despite using arguably the most biologically specific and severe morbidity associated with schistosomiasis. We also characterized the age-specific likelihood of periportal fibrosis, identifying a transitional age as young as 25 years. We identified future avenues for research into coinfections such as HIV and hepatitis B that appear to influence periportal fibrosis status even after controlling for a wide range of biosocial determinants of schistosome infection, treatment, and unrelated liver fibrosis. Future work is needed to understand if/how coinfections alter the pathogenesis of periportal fibrosis. Importantly, World Health Organization guidelines should be differentiated for schistosomiasis morbidities to discourage the use of infection status/intensity/prevalence as a proxy indicator for monitoring the elimination of periportal fibrosis as a public health problem.

## INTRODUCTION

Fifty-two countries require mass drug administration (MDA) for schistosomiasis with over 250 million people estimated to be infected and over 700 million people living in endemic areas^1^. The intestinal forms of chronic schistosomiasis, most prevalent in sub-Saharan Africa where *Schistosoma mansoni* is endemic, are characterised by periportal fibrosis (PPF).^1^ Mature schistosome flukes live in the mesenteric venules; paired female flukes release approximately 300-400 eggs per day.^2^ Only half of the eggs released successfully cross the intestinal mucosa to be excreted in stool. Eggs can become lodged in the gut wall or swept back into the vasculature of the liver and spleen. PPF is a chronic hepatic condition caused by an inflammatory response leading to granulomas and eventual fibrosis around parasite eggs in the portal veins and its segmental branches. The fibrosis can cause blockage of the main portal vein leading to portal hypertension and portosystemic collaterals such as oesophageal varices that when ruptured confer a high probability of death.^3^ Despite the clinical severity, there are no routine treatment strategies for individuals with PPF. The current World Health Organization (WHO) guidelines for schistosomiasis ^4^ focus on current infection as a proxy indicator for assessing morbidities associated with schistosomiasis, including PPF. Because of this focus, MDA with praziquantel to treat current infections only is the mainstay of treatment for schistosomiasis-associated conditions. However, there is no consensus as to the relevance of current schistosome infections for PPF.^5–8^

The epidemiology of PPF is poorly understood. In the context of mass drug administration and natural fluke death, PPF can outlast current schistosome infections.^6–9^ Existing studies have focused on unrepresentative samples of limited populations often targeting only adults.^5,6,10–12^ Information is lacking on the age-specific PPF likelihood over a cross-section of the endemic population currently eligible for praziquantel treatment (aged 5+ years). There are assumptions that the relationship of age with PPF is linear,^6,8,13^ despite intestinal schistosome prevalence being nonlinearly associated with age. Importantly, schistosome infections mostly have been studied in isolation for their associations to clinical liver pathologies,^6,8^ neglecting coinfections such as malaria or chronic hepatitis B/C and co-occurring conditions such as alcoholism which may confound or modify PPF.

We investigated PPF outcomes in a wide cross-section of age (5-90 years) for 2834 individuals in 38 rural communities within Western and Eastern Uganda. The aim of this study was to establish the epidemiology of PPF and the role of current individual schistosome infection status/intensity. We examined PPF likelihood over age, the contribution of current *S. mansoni* infection status to PPF, and the key biological and social risk factors associated to PPF with a particular focus on coinfections and comorbidities.

## Methods

### Participant sampling

This study was nested in the SchistoTrack Cohort, using the baseline data collection completed in January-February of 2022 in Buliisa, Pakwach, and Mayuge Districts in Uganda. MDA with praziquantel for individuals aged ≥5 years was started in 2003 with our districts receiving ≥13 rounds of treatment before this study with the most recent round in December 2020. Programmatic treatment coverage was on average 79·43% (std. dev. 21·93%), 80·18% (std. dev. 23·02%), and 77·74% (std. dev. 13·19%) in Buliisa, Pakwach, and Mayuge Districts, respectively. A total of 1459 households were randomly sampled using local registers from 38 villages. One child aged 5-17 years and one adult aged 18+ years were selected by the household head/wife and invited for the clinical assessments, while information also was collected on every member aged 1+ years from the sampled households.

Further details are provided in Supplementary Text S1. Figure 1 shows the participants sampled and the final number of 2834 individuals from 1442 households henceforth used for analysis. The number of individuals was calculated to detect a minimum effect size of ∼8% in an unevenly schistosome exposed/unexposed population (0·57) with a household design effect (1·136) and unbalanced strata/clusters at 97·5 power to account for multiple comparisons.

**Figure 1.**
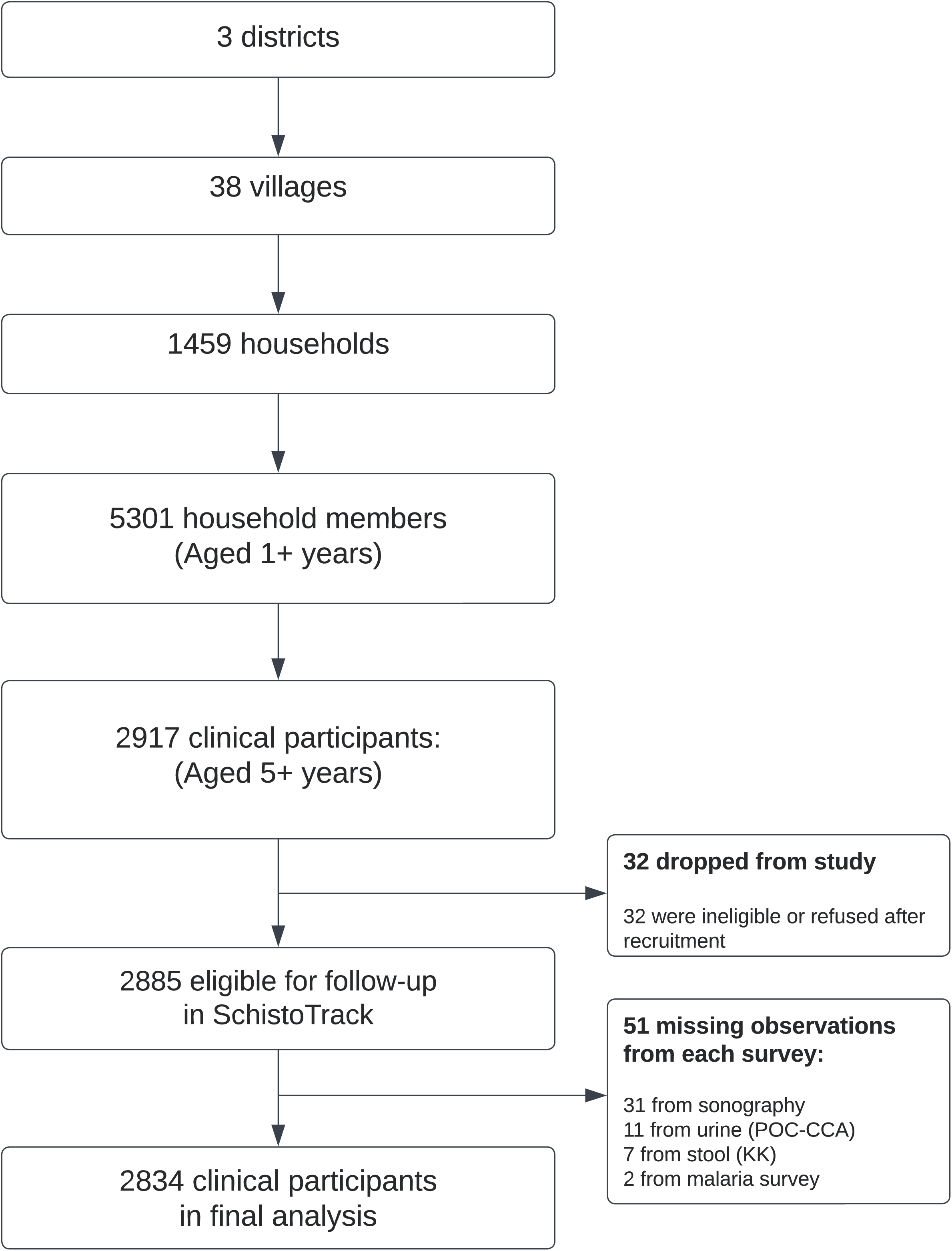
Participant sampling. The selection of study participants is shown and more fully described in the Methods section.

### Outcome

For PPF diagnosis, point-of-care B-mode ultrasound was used to acquire image patterns as described in the Niamey Protocol.^14^ Sonographers regardless of experience with the Niamey Protocol were retrained as part of the study. Inter-observer agreement among the eight study sonographers was evaluated to reach a consensus on abdominal sweep procedures for acquiring each PPF pattern. At least two sonographers were required to jointly scan and agree on grading. All observed patterns of PPF were recorded. The highest pattern assigned to each participant was used to construct a binary outcome where one or any of the C-F combinations was coded as one (PPF) and patterns A-B were coded as zero (no PPF). Philips Lumify C5-2 curved linear array transducers were used with the Philips Lumify Ultrasound Application v3·0 on Lenovo 8505-F tablets with Android 9 Pie. Lossless DICOM images and videos were saved for quality assurance.

### Exposures

*S. mansoni* infection was diagnosed using Kato-Katz (KK) microscopy ^15^ and point-of-care circulating cathodic antigen tests (POC-CCA; batch 210811080).^16^ For KK, a single stool sample, double slide thick smears were prepared and read by two technicians with the results from the 41·7mg slides averaged and multiplied by 24 to calculate eggs per gram (EPG). KK infection status was positive when EPG ≥1. WHO infection intensity categories were constructed with none, low, moderate, and heavy as 0, 1-99, 100-399, 400+ EPG respectively [8]. POC-CCA results were interpreted as negative, trace (barely visible test line), positive 1 (test line fainter than control line), positive 2 (test line similar to control line) and positive 3 (test line much darker than control line). Infection status by POC-CCA was positive if positive 1-3 was assigned; two alternative variables were constructed with trace positive (Tr+) and negative (Tr-). Ten percent re-readings of participant diagnoses by a senior technician were completed for KK slides and images of POC-CCA. Village-level variables were constructed from individual infection indicators (Supplementary Table S1).

### Covariates

Detailed definitions are provided in Supplementary Text S1. At the individual level, we measured age, gender, religion, tribe, water contact, education, and occupation. The history of liver diseases, human immunodeficiency virus (HIV+) status, previous use of praziquantel or antiretroviral therapy (ART), complicated/uncomplicated malaria, hepatitis B/C, alcohol use, and smoking were recorded. Ultrasound-based first-pass assessments of ascites, chronic hepatitis/early cirrhosis-like disease, and fatty-like livers were completed, and rapid diagnostic tests of malaria were conducted (SD Bioline Malaria Ag P.f/Pan). At the household level, variables were constructed for total years of settlement in the village, home quality score, electricity availability, social status, home ownership, and improved drinking water, hygiene, and sanitation. Other variables included water body types in the village of residence, availability of at least one public latrine within the village, and district.

### Statistical analyses

All the analyses were conducted with R v4·1. The functional form of the relationship between age and PPF was identified using the locally weighted scatterplot smoothing (LOWESS) and Generalised Addictive Models (GAM) where spans were chosen through 10- fold cross-validation (CV).^17^ To identify key transition points in PPF likelihood across age, an alternative method of free-knot spline modelling framework with generalised CV was used.^18^

The intraclass correlation coefficients (ICC) in an empty multi-level logistic model was used to examine the maximum clustering of PPF within households and villages.^19^ A Hausman test ^20^ was used to compare the empty multilevel model with an empty logistic model to investigate village fixed versus random effects. PPF predictors were chosen with likelihood ratio tests (LRTs) of univariate models when p-values were ≤0·05. ART and other variables with ≤1% frequency was dropped (Table 1). To account for the sampling design, fully adjusted multivariable logistic models were run with clustered standard errors at the household level.^21^ Covariates selected by LRTs but with variance inflation factor (VIF) >10 were removed and presented in alternative models. The model building procedure was rerun for subgroups of only children, adults, and individuals from Pakwach District. Participants with both PPF, history of liver diseases, chronic hepatitis/early cirrhosis-like disease, or fatty-like livers (Niamey Protocol XY patterns) were recoded as zero (no PPF) and models were rerun. To assess the predictive capacity of models, 10-fold CV was used with 5-fold CV for sub-group analyses with fewer observations.^22^

**Table 1.**
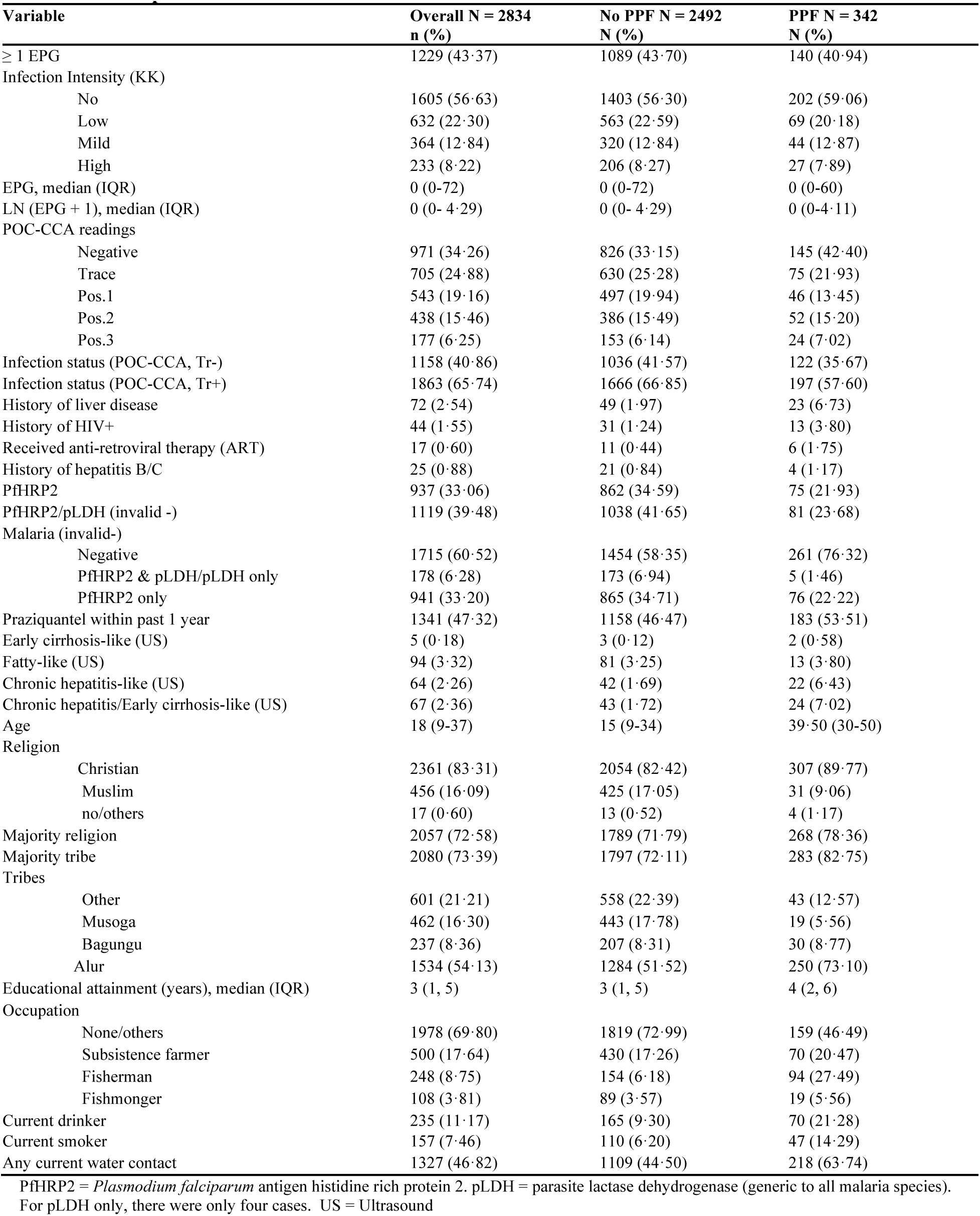
Participant characteristics.

### Ethics approvals

Data collection and use were reviewed and approved by Oxford Tropical Research Ethics Committee (OxTREC 509-21), Vector Control Division Research Ethics Committee of the Uganda Ministry of Health (VCDREC146), and Uganda National Council of Science and Technology (UNCST HS 1664ES).

## RESULTS

### PPF and *S. mansoni* prevalence

Participant characteristics are shown in Table 1 with liver fibrosis patterns demonstrated in Figure 2. PPF prevalence is shown in Figure 3a-d. Household and village covariates are presented in Supplementary Tables S2. Over 12% (344/2834) of participants were diagnosed with PPF; C image patterns were most common (5·39%, 153/2834). The highest prevalence of PPF at 19·46% was found in Pakwach (180/925), followed by 12·11% (115/950) in Buliisa and 4·90% (47/959) in Mayuge District. Across all individuals, PPF prevalence was higher among males when compared to females, and among adults when compared to children. Only 5 of 2834 (0·18%) participants presented with any grade of ascites; all ascites cases had PPF. There were 2·34% (69/2834) and 3·32% (94/2834) of participants, respectively, with chronic hepatitis/early cirrhosis-like or fatty-like livers detected by ultrasound. Over 7% (24/342) participants with PPF had a chronic hepatitis/early cirrhosis-like liver. By KK, 43·37% (1229/2834) of individuals were infected, with 8·22% (233/2834) heavily infected. POC-CCA Tr+ at 65·73% (1863/2834) was higher than KK prevalence and POC-CCA Tr- at 40·86% (1158/2834) was similar to KK prevalence.

**Figure 2.**
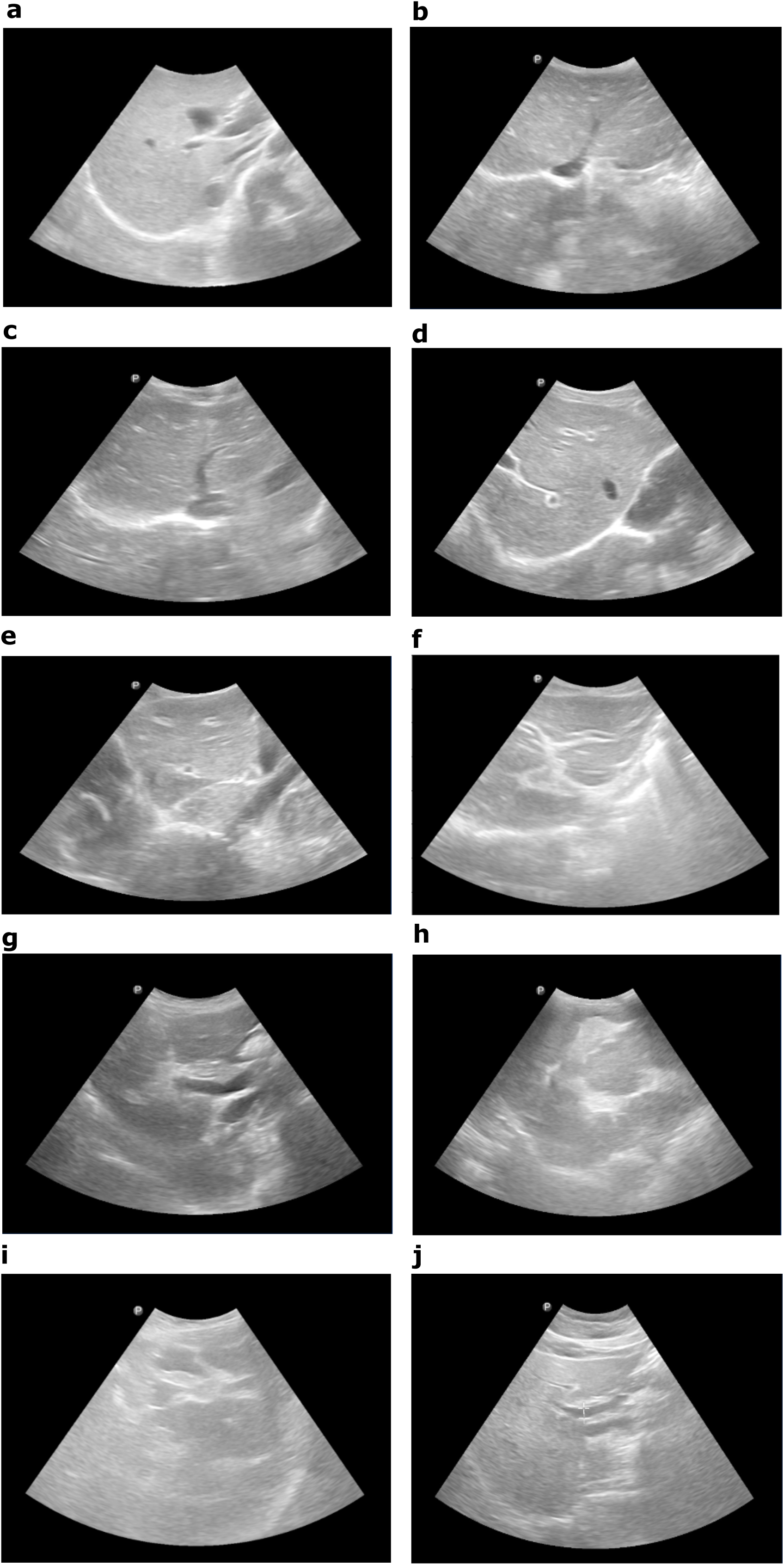
Periportal fibrosis image patterns and comorbidities. Patterns as described in the Niamey Protocol are shown and suspected diffuse liver diseases such as chronic hepatitis/early cirrhosis-like livers or fatty-like livers are noted where observed. **a.** A normal liver pattern is shown with no comorbidities. **b.** A B1 image pattern depicting flying saucers or starry sky is shown. **c.** A B2 spider thickening pattern is shown. **d.** A C1 image pattern (prominent peripheral rings) with a chronic hepatitis-like liver is shown. The diffuse disease was suspected to be more hepatitis-like in pattern than cirrhosis-like due to the retained liver surface regularity. **e.** A C1 image pattern is shown with a more cirrhosis-like, possibly advanced cirrhosis, comorbidity where there was evidence of a possible shrunken liver and gross surface irregularity. **f.** A C2 (prominent pipe stems) image pattern is shown. **g.** A D image pattern (ruff portal bifurcation) is shown. **h.** Image Pattern E (patches representing occluded, bright white vessels) is shown. **i.** The most severe pattern of PPF (F, a bird’s claw) is shown. **j.** A fatty-like liver with no evidence of image patterns B-F is shown.

**Figure 3.**
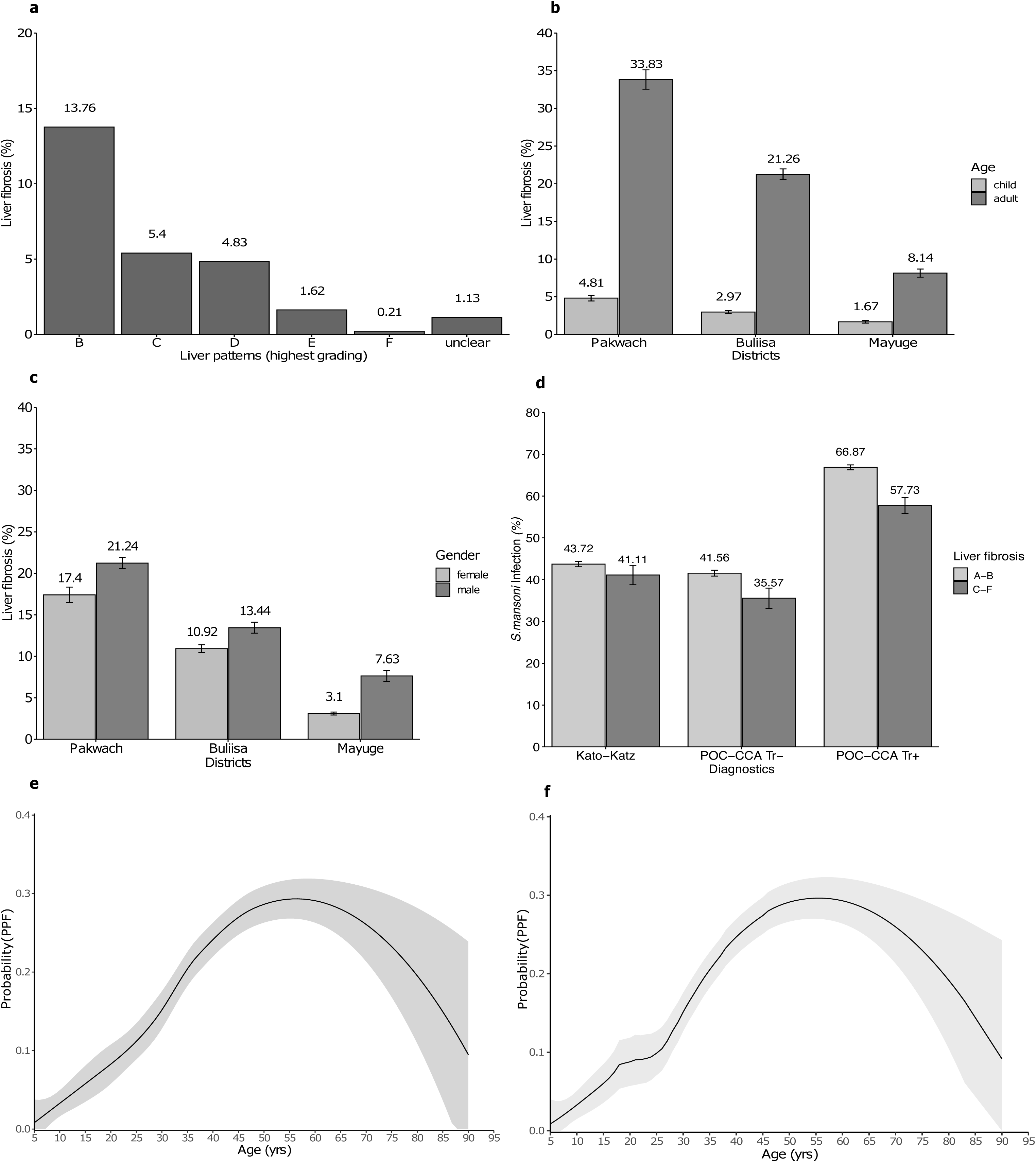
Prevalence of periportal fibrosis and age-specific likelihood. N=2834. Pakwach N = 925. Buliisa N = 950, Mayuge N= 959. **a.** The prevalence of liver patterns is shown, whereby only the highest pattern per participant was included. **b.** Prevalence of periportal fibrosis (liver pattern C-F) by study districts and age group is shown. **c.** Prevalence of periportal fibrosis by study district and gender is shown. **d**. Prevalence of *S. mansoni* infection by periportal fibrosis is shown. **e.** Locally weighted scatterplot smoothing (LOWESS) was applied. The panel shows fewer data points among the study sample between the ages of 16-19 years. The predicted probability of periportal fibrosis at each value of age using the LOWESS default span (0·75) is shown. **f**. Similar to panel e, the predicted probability of periportal fibrosis is shown at each value of age, but differently here using a span of 0·62 selected by 10-fold CV.

### Age-specific PPF

The study participants had a median age of 18 years (IQR 9-37) with a range of 5-90 years. Figure 3e-f presents the functional form of age with PPF. Age showed a linear relationship from childhood through to adolescence (5-25 years) with the likelihood of PPF changing exponentially in young adulthood to middle age (26-45 years). The likelihood of PPF becomes steady until around 55-60 years of age where there is a sharp decline in the likelihood of PPF approaching <10%. A transitional point in PPF likelihood was observed around age 25-26 years (Supplementary Figure S1). Both LOWESS and GAM analyses suggested non-monotonic relationships with an approximately quadratic relationship (inverse-U) of age with PPF.

### Determinants of PPF

No intraclass clustering of PPF was observed within households, whereas 14% of variation was found within villages. Figures 4-5 show unadjusted associations. In fully adjusted models (Figure 6 with alternative infection indicators in Supplementary Figure S2), no individual-level *S. mansoni* infection indicators were associated with PPF. This result was robust to sub-group analyses for children, adults, Pakwach District only, and recoding of comorbid diffuse liver disease (XY patterns) as no PPF (Supplementary Figures S4-S7). Each one-year increase in age was associated with a 14·3% increase in the likelihood of PPF (95% CI 1·10-1·18). The quadratic term for age that captured the inverse-U relationship showed a negative association with PPF, although this effect was small <0·001% (95% CI 0·998- 0·999). Female participants were 32·2% less likely than males to have PPF (95% CI 0·49- 0·94). Fishermen were 71·2% more likely than unemployed or other occupations to have PPF (95% CI 1·12-2·63). Subsistence farmers were 32·6% less likely to have PPF when compared to unemployed or other occupations (0·47, 0·98). Participants with a reported previous diagnosis of liver diseases and HIV+ by a government health worker had 2·19 and 2·31-fold increased likelihood of PPF, respectively. Participants with ultrasound-detected chronic hepatitis/early cirrhosis-like disease had a 2·84-fold higher likelihood of PPF (95% CI 1·42-5·67) than individuals without these ultrasound-detected diffuse liver diseases. No household-level factors were significantly associated with PPF.

**Figure 4.**
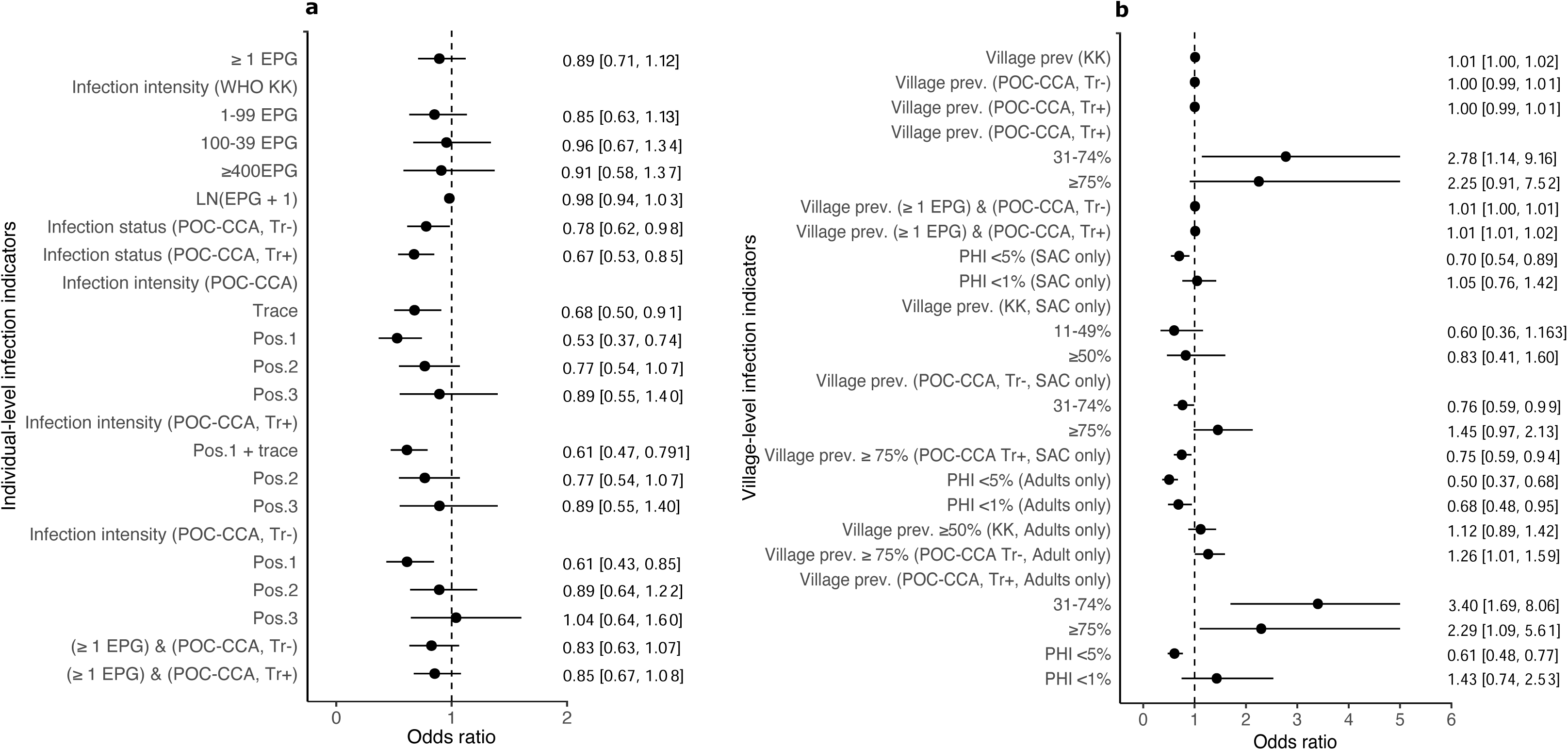
Unadjusted models for *S. mansoni* infections and periportal fibrosis. Odds ratios (ORs) and corresponding 95% confidence interval (CI) represent univariate logistic regression models. **a.** Unadjusted relationships are shown between periportal fibrosis and sociodemographic indicators. **b**. Unadjusted relationships are shown between periportal fibrosis and morbidity indicators.

**Figure 5.**
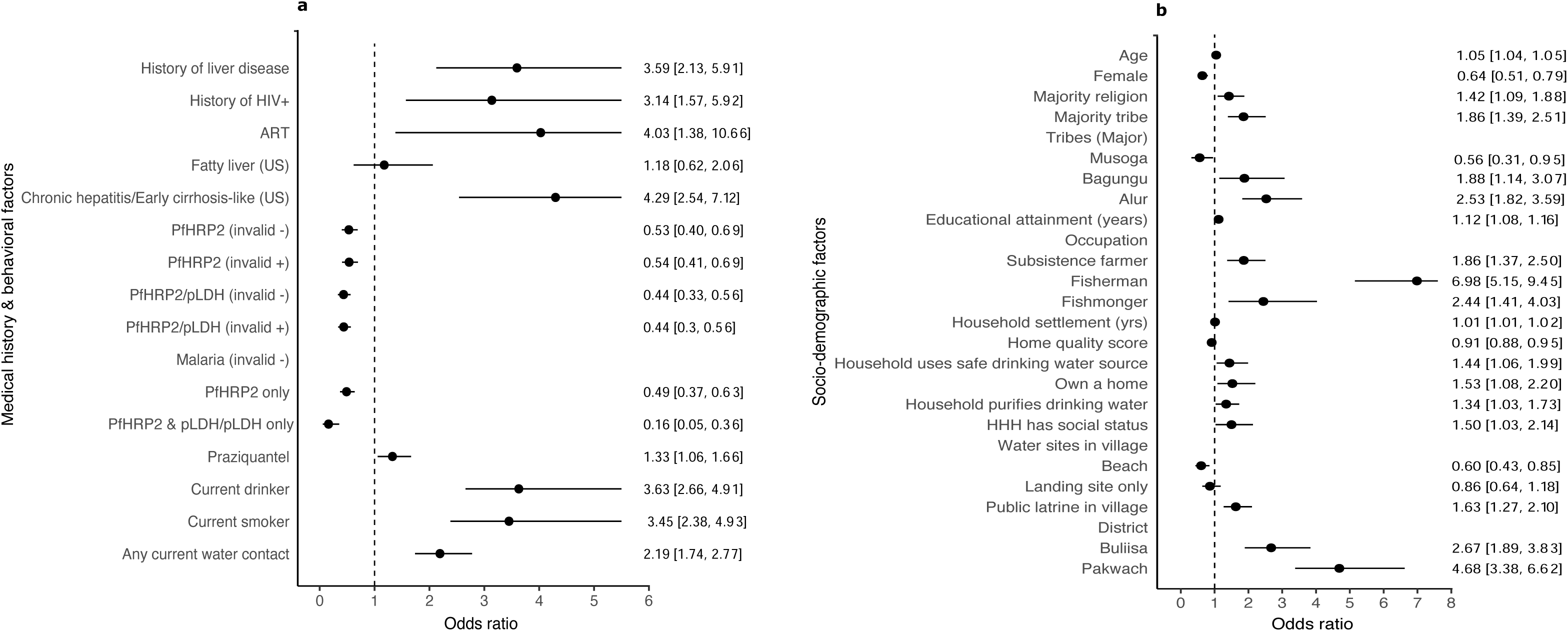
Unadjusted models for covariates and periportal fibrosis. Odds ratios (ORs) and corresponding 95% confidence interval (CI) represent univariate logistic regression models. PfHRP2 = *Plasmodium falciparum* antigen histidine rich protein 2. pLDH = parasite lactase dehydrogenase (generic to all malaria species). For pLDH only, there were only four cases. US = Ultrasound. **a**. Unadjusted relationships are shown between PPF and coinfections as well as other medical history factors. Alcohol and smoking variables were run for adult only models (individuals aged 18+ years). **b.** Unadjusted relationships are shown between PPF and sociodemographic and ecological indicators.

**Figure 6.**
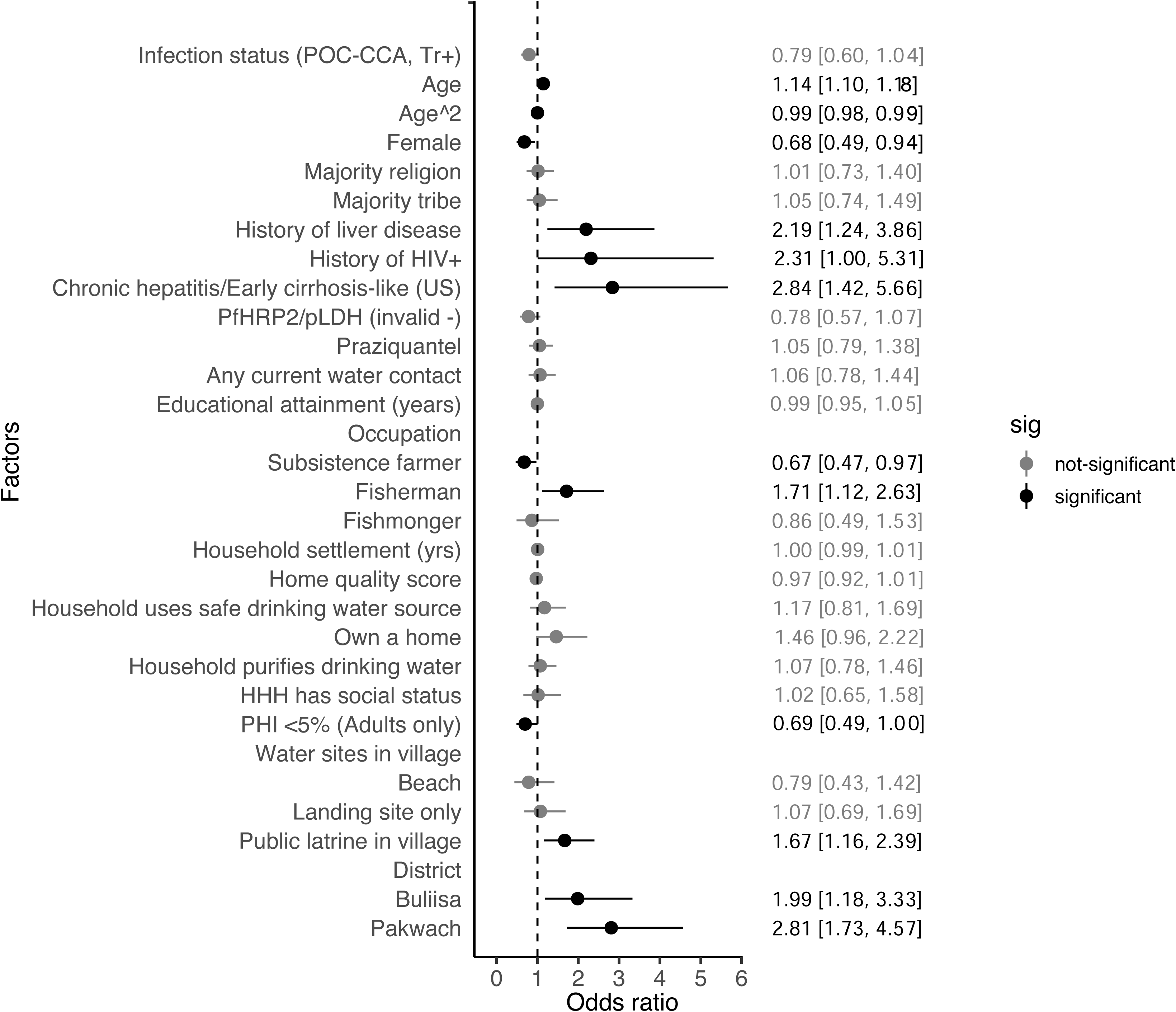
Determinants of periportal fibrosis. A fully adjusted model is shown with variables selected through LRT with p-value ≤0·05. Black represents significant relationships of p-value ≤0·.05. No variables in the model had a VIF >10. The mean area under the ROC (by 10-fold CV) was 0·82. Hausman test result of an empty random vs village fixed effects model (with 38 villages) was chi^2^= 3·90, p-value<0·05. PfHRP2 = *Plasmodium falciparum* antigen histidine rich protein 2. pLDH = parasite lactase dehydrogenase (generic to all malaria species). For pLDH only, there were only four cases. US = Ultrasound.

Village and district characteristics were associated with PPF (Figure 6). Living in a village with <5% prevalence of heavy infection intensity (PHI) among adults was borderline significantly associated with a 30·2% reduced likelihood of individual PPF (95% CI 0·48- 1·00). Living in a village with ≥1 public latrine was associated with a 66·8% higher likelihood of individual PPF (95% CI 1·16-2·40). Only 1/3^rd^ or less of the public latrines across the villages were reported to be in use at the time of the survey due to flooding.

Participants living in Buliisa had 1·99 and in Pakwach had 2·81-fold higher likelihood of PPF than participants living in Mayuge District. When exact tribe was investigated (Supplementary Figure S3), participants from the Alur tribe were found to be 78·3% more likely to have PPF when compared to other tribes. Alur and Bagungu tribes were similar with respect to the percentage of fishermen (10·89%; 167/1534 vs. 10·97%; 26/237), whereas the Musoga tribe had fewer fishermen (4·33%; 20/462). The tribes of Alur, Bagungu, and Musoga were highly collinear with district effects (VIF>10). All models showed high predictive accuracy of PPF with 10-fold CV area under the receiver operating characteristic (ROC) curve of >80%.

## DISCUSSION

There are no guidelines within the recent 2020-2030 WHO Roadmap for Neglected Tropical Diseases ^23^ for routine surveillance and treatment of morbidity associated with schistosomiasis. Here we showed that severe morbidity caused by *S. mansoni* persisted in areas that have received annual community-wide MDA. We conducted a population-based study of 2834 individuals aged 5-90 years across 38 rural villages in diverse regions of Uganda. Our findings showed that age-specific PPF likelihood was nonlinear, drastically changing around age 25 years. Underlying liver diseases associated with the history of HIV coinfection and ultrasound-detected chronic hepatitis/early cirrhosis-like livers as opposed to current singular *S. mansoni* infections were key predictors of having PPF.

Age-specific PPF likelihood was nonlinear, suggesting the association of age with PPF changes over the life course of an individual. The trends found for PPF likelihood do not agree with well-established age-specific infection curves.^24^ Age-specific infection prevalence and mean intensity are known to peak in school-aged children and decline sharply in adults, which has provided past evidence for acquired immunity even in the context of steady infection exposure.^25^ Yet, PPF likelihood appears to linearly increase with age in children whilst exponentially increasing in young adults. There appeared to be a proximal transition age of 25 years in likelihood of PPF before levelling off in older adults and sharply declining in the eldest individuals. These results suggest that other factors unrelated to cumulative history of schistosome infection are co-occurring during early adulthood and influencing PPF onset or progression. The drastic decline in older age, beginning from age 55-60, may indicate excess mortality due to *S. mansoni-*associated morbidities in our study areas as the average life expectancy in rural Uganda is 65 years.^26^ Future work is ongoing in SchistoTrack to identify prospectively the risk factors that individuals encounter over their life course, the onset of PPF, and the likelihood of death due to PPF.

There was no consistent association of individual *S. mansoni* infection status/intensity or village-level prevalence irrespective of diagnosis with KK microscopy or POC-CCA in either unadjusted or adjusted regressions. This lack of association was robust even when examining only adults. Furthermore, recent receipt of praziquantel was not correlated with PPF likelihood suggesting current treatment through MDA which has been shown to approximate past treatment likelihood ^27^ may not address severe *S. mansoni* morbidity. This finding agrees with a meta-analysis by Andrade et al. showing morbidities associated with *S. mansoni* have poorer resolution rates after MDA as compared to *S. haematobium.*^28^ FibroSchot is an ongoing trial in Western Uganda that is causally assessing the potential of praziquantel outside of MDA to prevent and reverse PPF.^29^ The long history of MDA (≥13 years) in our study areas coupled with high reported programmatic coverage (>75%) in individuals aged ≥5 years and well-demonstrated poor cure rates with praziquantel further suggest that blanket treatment campaigns cannot geographically eliminate PPF. ^27,30^

Infection prevalence and intensities in school-aged children were poor proxy indicators of the likelihood of an individual—whether another child or adult—having PPF. However, the PHI in adults was suggestive of a borderline positive association with PPF. Future work might investigate revising WHO proxy indicators. Village infection prevalence, including groupings of WHO categories used to guide the frequency of mass drug administration, were not consistently associated with PPF likelihood.^4^ This result may indicate that current guidance for treating infections through MDA cannot be used for hotspot identification of likely areas to have residual PPF. Any level of village infection prevalence could correlate with PPF likelihood. However, the persistence of heavy intensity infections in only adults could be investigated in future work as an indicator of community transmission intensity and potentially used for PPF hotspot identification. Given that acquired immunity to reinfection in adults for *S. mansoni* has been well demonstrated.^25^ the persistence of heavy intensity infections in adults might reflect an excessively high historic force-of-infection.

PPF shares some epidemiology with *S. mansoni* infection yet has little overlap with predictors of praziquantel receipt. Age, gender, and being a fisherman were found to be significant predictors of PPF and in the same directions as expected for predicting infection likelihood.^24^ Yet, the effect sizes were approximately 1/4^th^ or less the size of the effects for HIV+ history, chronic-hepatitis/early cirrhosis-like disease, and study district. Being a fisherman is related to water contact and, in turn, infection or reinfection risk.^24^ However, it is unclear whether being female confers some protection against PPF due to water contact behaviours or differences to health care access. Women have been shown to access health care more routinely within their reproductive years ^31^ and signs of liver fibrosis may be picked up at an early stage. For example, any liver enlargement, tenderness, or changes in consistency might be found incidentally during abdominal palpations for pregnancy which then warrant further examinations of the liver and lead to care. Key praziquantel treatment predictors including social status, home quality (wealth), education, and home ownership, among other factors were not relevant for predicting PPF.^27^ This finding further supports that both direct and indirect measurements of past praziquantel treatment show no correlation with PPF.

Commonly assessed co-occurring conditions such as alcohol use and malaria were not associated with PPF, despite correlations with PPF in other studies ^32,33^ that did not adjust for diffuse liver diseases or HIV+ history. It is possible the correlation of alcoholism with PPF in adults was not detectable in our study if it manifested in older individuals with advanced cirrhosis where our data may have exhibited survivor bias associated with PPF.^34^ The lack of association of current drinking with PPF was not due to accounting for chronic hepatitis/early cirrhosis-like disease; when this variable was removed from the model, current drinking remained insignificant. For the malaria insignificance for PPF, the parasite predominantly affects the spleen. The liver is only briefly involved in the life cycle where sporozoites mature into schizonts, especially for *P. falciparum* which was most common in our study areas. The peak of malaria prevalence and its associated severe morbidities often are amenable to antimalarials and found in children <5 years of age who were not studied here.

History of HIV+ and ultrasound-detected chronic hepatitis/early cirrhosis-like disease were positively correlated with PPF. Each comorbidity has been shown previously to singularly contribute to the likelihood of PPF.^7,35^ HIV+ has been shown to correlate with hyaluronic acid levels,^36^ which are indicative of liver fibrosis. What remains unclear is the mechanism of HIV+ and *S. mansoni* interactions on the ability of the host to mount a granulomatous response to eggs or for the associations with prognostic outcomes such as severe anaemia or portal hypertension. For HIV+, it is possible that status was underestimated if there were individuals who have yet to be diagnosed or who had concerns in reporting due to community stigmatisation. Chronic hepatitis or cirrhosis causes diffuse liver pathologies affecting liver functionality. Confirmatory testing that considers viral hepatitis diagnostics, alternative imaging modalities, and fibrosis scores is needed in future work for distinguishing and staging hepatitis from cirrhosis. Considering causes of hepatitis-like livers, hepatitis C is rare in our study areas, whereas hepatitis B may be a promising line of investigation as it has been shown to exceed 10% in adults in Western Uganda,^37^ and will be incorporated in future SchistoTrack follow-ups. Confounding by aflatoxins also must be investigated, which is challenging given the diverse subtypes and sampling frames of either human or grain.

Importantly, for future investigations of diffuse liver disease interactions with PPF, an important consideration is to separate image pattern outcomes. A PPF pattern of D is less diffuse than other PPF patterns; hence, viral hepatitis coinfections might be suspected to only correlate with specific image patterns.

We found that individuals in Pakwach and Buliisa Districts were more likely to have PPF than individuals in Mayuge District. Geographical differences in PPF have been documented previously between Western and Eastern Uganda.^13,38^ Our results suggest the district effects are not due to environmental conditions and, in turn, possible differences in parasite strains. No ecological predictors were correlated with PPF. Moreover, individual tribes were highly collinear with district effects. Although there was no household clustering of PPF, we are unable to make any inferences on the relevance of genetic ties for PPF as we did not require the sampled adult to be the parent of the sampled child. To identify any shared genetic effects through tribe, there is a need to prospectively examine only child-adult or sibling pairs.

Research is ongoing in SchistoTrack to assess the development of PPF across a five-year period in children belonging to households that have parents with PPF. Alternatively, the effect of tribe might represent differences in water contact behaviours not captured through differences in the prevalence of fishermen or differences in access to health care.^39,40^

Indicators for current individual infection status/intensity were not associated with the likelihood of PPF. Proxy indicators for monitoring morbidity should be tailored to the epidemiology of the morbidity. It is well-established that PPF is more prevalent in adults. PHI in adults, rather than the current WHO guideline noting PHI in school-aged children, might be a better indicator to assess progress towards control of morbidities associated with *S. mansoni*. Programmes aimed at controlling PPF should consider an integrated approach, especially for adults in endemic settings. Improved access to health care services for the treatment of schistosomiasis and capacity building within local health systems for recognising PPF are needed. Multimorbidity could be addressed through case management strategies to explore early identification of PPF. Areas where integration across diseases might be explored may include investigating how hepatitis B/C and HIV clinics could be expanded to include diagnosis for schistosomiasis-associated morbidity. The results of this study, if replicated in other settings, could provide interventions to be tested in future randomised-controlled trials for improving morbidity management for severe hepatic schistosomiasis.

## Author contributions

Conceptualization: GFC and NBK. Data curation and validation: SA. Formal analysis: SA. Investigation, methodology, validation, visualization, and writing – original draft: SA and GFC. Writing – review and editing: SA, BN, SM, BK, CKO, NBK, and GFC. Funding acquisition and supervision: GFC. Resources: BN, GFC, and NBK. Data collection: BN, SM, BK, NBK, and GFC.

## Data sharing

Data is not publicly available due to data protection and ethics restrictions related to the ongoing nature of the SchistoTrack Cohort and easily identifiable nature of the participants.

## Acknowledgements

This work was supported by the Nuffield Department of Population Health Pump Priming Fund, Wellcome Trust Institutional Strategic Support Fund (204826/Z/16/Z), John Fell Fund, Robertson Foundation Fellowship, and UKRI EPSRC Award (EP/X021793/1) to G.F. Chami. We are thankful for the involvement and trust from our study participants. There were extensive SchistoTrack field teams involved including malacologists, surveyors, nurses, sonographers, and laboratory technicians. Without the support of the Uganda Ministry of Health, local district leaders, focal health workers, and village health teams, we could not have worked so closely in partnership with the study communities. We also have many thanks to give to the Oxford team for support with fieldwork, data wrangling, and feedback during group meetings.

## Supplementary information

### Text S1 Variable definitions

#### Individual-level variables

Age was a continuous variable to the nearest year. Age squared was incorporated in models to account for nonlinearity with PPF. Gender and religion were defined as binary variables with females and Muslims coded as one, respectively. The majority religion was a binary variable, which was coded as one if participants belonged to the majority religion within their own village.

Medical history was recorded by household surveyors who used modified versions of the World Health Surveys to collect information on past diagnoses.^1^ Individuals were asked whether a government health worker had diagnosed them with a set of conditions. Each condition was constructed as a binary variable. The conditions asked about included liver diseases (cirrhosis, scarring or fibrosis), HIV, and hepatitis B/C. The WHO Stepwise approach for risk factor surveillance (STEPS) was used to gather information on non-communicable diseases such as smoking and alcohol drinkers.^2^ Individuals were coded as current smokers if any tobacco products were used within the past 12 months preceding the study. Individuals were coded as current drinkers if they had consumed any alcohol within the past 12 months. Only study participants aged 10+ years were surveyed on alcohol and smoking behaviours; therefore, these variables were only used in the adult sub-group model.

Previous treatment with praziquantel was coded as a binary variable where participants who received or ingested praziquantel in the past one year were coded as one. Mass drug administration began in all study districts in 2003. Buliisa and Pakwach Districts received 13 annual rounds of mass drug administration, whereas Mayuge District had received 15 rounds of mass drug administration preceding the SchistoTrack baseline. There was no mass drug administration the year before the survey due to COVID-19 lockdowns, so receipt of praziquantel would have been through other means such as leftover medicines remaining with community medicine distributors. The last round of mass drug administration in all our study districts was conducted in December 2020, using both school-based and community-based distribution. Current anti-retroviral therapy was also coded as one if participants confirm they have taken western medicine in the past one month and anti- retroviral therapy was indicated as one of them.

Ultrasound was used to gather information on suspected current diffuse liver diseases. Chronic hepatitis-like, cirrhosis-like, and fatty-like livers were noted.^3^ Chronic hepatitis-like livers were observed with diffuse, inhomogeneous echogenic nodules and no surface (liver outline) irregularities. Cirrhosis-like livers were defined as having a coarse liver parenchymal echotexture, rounded/blunted caudal liver edge, and potential surface irregularities. Possible shrunken livers or reduced left and right medial sections were considered albeit not required for our definitions of cirrhosis-like livers as we did not only record advanced cases. As chronic hepatitis can mimic early/compensated cirrhosis when diagnosed only via ultrasound,^4^ a binary variable equal to one was generated if there was any evidence of either chronic hepatitis-like or cirrhosis-like livers. In our data, all five cases of cirrhosis-like livers had sharp caudal edges suggesting early cirrhosis. Only one of five cases of cirrhosis-like livers had any (here, slightly serrated) surface irregularities. As we were interested in chronic underlying liver diseases, separate ‘starry-like’ patterns for acute hepatitis were not investigated outside the image pattern B provided in the Niamey Protocol. Fatty-like livers were defined as having diffuse parenchymal brightness with observed liver-to-kidney contrast and posterior attenuation. B-mode ultrasound has shown to be reliable for steatosis exceeding 20%,^5^ hence we were likely to miss mild fatty-like liver cases. Ascites was noted when fluid build-up was observed in any of the four abdominal quadrants. If fluid was observed, but not present in all four quadrants then it was graded as mild. If fluid was observed in all four quadrants, but the abdomen and organs were still clearly distinguishable then ascites was graded as moderate. When there was fluid build-up in all four quadrants to the point that the quadrants were indistinguishable, and organs were difficult to differentiate then ascites was graded as severe.

Any water contact was coded as one if a participant did at least one of following activities in the lake or river near their village including fishing, fishmongering, collecting papyrus/shells, retrieving drinking water, washing clothes with/without soap, washing jerry cans or other household supplies, swimming, or playing in water. Education was defined as a count variable from 1-14, whereas 1-7 represented primary school years, 8-13 captured lower and upper secondary school and 14 represented participants that have completed a post-secondary education. Education also was expressed as a categorical variable of no education, primary education, and secondary education or higher. Occupation was defined as a categorical variable of subsistence farmer, fishermen, fishmongers and no and other occupations as reference category.

Household-level variables including water, sanitation, and hygiene indicators, social status, home quality, electricity, ownership, and number of rooms as well as village ecology and infrastructure were defined as described in Chami et al. 2016.^6^ The years the household had settled in the village was a continuous variable defined as the years anyone in the household or an immediate relative of the household head (who was in the same household) had been living in the village. Outliers/typos in this variable were recoded to the next nearest value. The recoded outliers were -12 and 3333501, which were recoded to 12 and 50, respectively.

**Table S1.** *S. mansoni* infection indicators and their alternative definitions.

| **Infection Indicator** | **Definition** |
| --- | --- |
| LN(EPG + 1) | Natural log of average egg per gram of stool (+1) |
| Infection intensity (POC-CCA, Tr+) | Infection categories from urine sample using point-of-care circulating cathodic antigen (POC-CCA) test; trace positive |
| Infection intensity (POC-CCA, Tr-) | Infection categories from urine sample using POC-CCA test; trace negative |
| Village prev. (KK) | Village level prevalence of infection by KK |
| Village prev. $\geq 50\%$ (KK) | Village prevalence setting of schistosomiasis infection by KK. |
| Village prev. (POC-CCA, Tr-) | Village level prevalence of infection by POC-CCA trace negative. |
| Village prev. (POC-CCA, Tr+) | Village level prevalence of infection by POC-CCA trace positive; continuous |
| Village prev. $\geq 31\%$ (POC-CCA, Tr-) | Village prevalence setting of schistosomiasis infection by POC-CCA trace negative. |
| Village prev. (POC-CCA, Tr+) | Village prevalence for *S. mansoni* infection by POC-CCA trace positive. |
| Village prev. ($\geq 1$ EPG) & (POC-CCA, Tr-) | Village-level infection prevalence by agreement of KK and POC-CCA trace negative |
| Village prev. ($\geq 1$ EPG) & (POC-CCA, Tr+) | Village-level infection prevalence by agreement of KK and POC-CCA trace positive |
| Village prev. (KK, SAC only) | Village prevalence of infection (KK) among school-age children |
| Village prev. (POC-CCA, Tr-, SAC only) | Village prevalence of infection (POC-CCA trace negative) among school-age children |
| Village prev. (POC-CCA, Tr+, SAC only) | Village prevalence of infection (POC-CCA trace positive) among school-age children |
| Village prev. (KK, Adults only) | Village prevalence of infection (KK) in adults only |
| Village prev. (POC-CCA, Tr+, Adults only) | Village prevalence of infection (POC-CCA trace positive) among adults |
| Village prev. (POC-CCA, Tr-, Adults only) | Village prevalence of infection (POC-CCA trace negative) among adults |
| PHI $< 5\%$ | Study participant lived in village with prevalence of heavy infection intensity (PHI; 400+ eggs per gram (EPG)) $< 5\%$ among school-aged children (SAC) and adults |
| PHI $< 1\%$ | Study participants lived in village with PHI $< 1\%$ among SAC and adults |
| PHI $< 5\%$ (SAC only) | Study participants lived in village with PHI $< 5\%$ among SAC only |
| PHI $< 1\%$ (SAC only) | Study participants lived in village with PHI $< 1\%$ among SAC only |
| PHI $< 5\%$ (Adults only) | Study participants lived in village with PHI $< 5\%$ among adults only |
| PHI $< 1\%$ (Adults only) | Study participant lived in village with PHI $< 1\%$ among adults only |

**Table S2.**
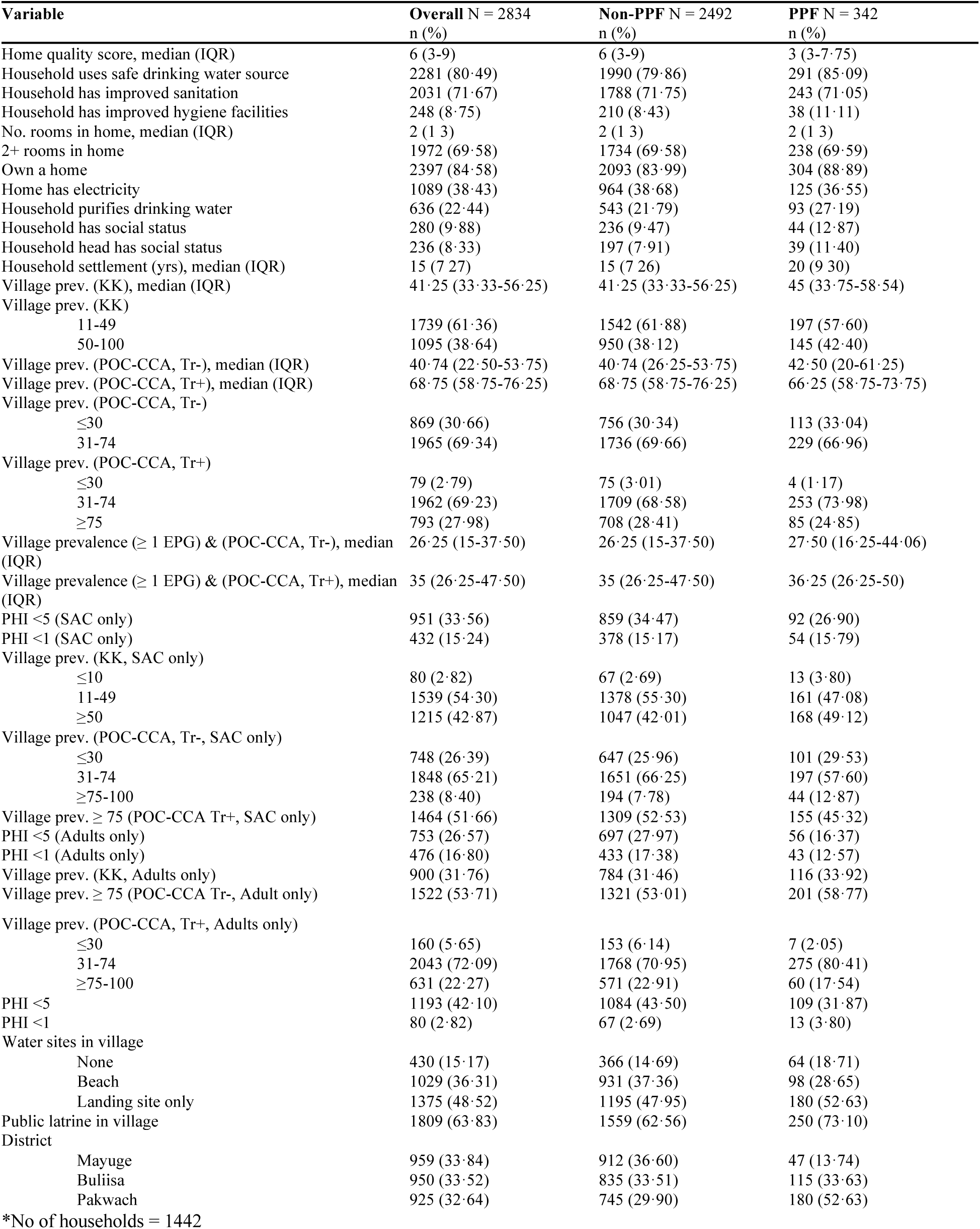
Descriptive statistics of household and village-level variables.

**Table S3.** Descriptive statistics of villages-level infection indicators.

| Variable | N | n (%) |
| --- | --- | --- |
| Village prev. (KK), median (IQR) | 38 | 41.25 (33.44-55.94) |
| Village prev. (KK) | 38 |  |
| 11-49% |  | 24.00 (63.16) |
| ≥ 50% |  | 14.00 (36.84) |
| Village prev. (POC-CCA, Tr-), median (IQR) | 38 | 40.74 (26.88, 53.44) |
| Village prev. (POC-CCA, Tr+), median (IQR) | 38 | 68.12 (59.06-75.94) |
| Village prev. ≥31 (POC-CCA, Tr-) | 38 |  |
| ≤ 30% |  | 12.00 (31.58) |
| 31-74% |  | 26.00 (68.42) |
| Village prev. (POC-CCA, Tr+) | 38 |  |
| ≤ 30% |  | 1.00 (2.63) |
| 31-74% |  | 26.00 (68.42) |
| ≥ 75-100% |  | 11.00 (28.95) |
| Village prevalence (≥ 1 EPG) & (POC-CCA, Tr-), median (IQR) | 38 | 25.62 (16.88- 37.50) |
| Village prevalence (≥ 1 EPG) & (POC-CCA, Tr+), median (IQR) | 38 | 35.62 (26.56-46.88) |
| PHI <5% (SAC only) | 38 | 14.00 (36.84) |
| PHI <1% (SAC only) | 38 | 7.00 (18.42) |
| Village prev. (KK, SAC only) | 38 |  |
| ≤ 10% |  | 1.00 (2.63) |
| 11-49% |  | 21.00 (55.26) |
| ≥ 50% |  | 16.00 (42.11) |
| Village prev. (POC-CCA, Tr-, SAC only) | 38 |  |
| ≤ 30% |  | 10.00 (26.32) |
| 31-74% |  | 25.00 (65.79) |
| ≥ 75% |  | 3.00 (7.89) |
| PHI <5% (Adults only) | 38 | 10.00 (26.32) |
| PHI <1% (Adults only) | 38 | 6.00 (15.79) |
| Village prev. (KK, Adults only) | 38 | 12.00 (31.58) |
| Village prev. ≥ 75 (POC-CCA Tr+, SAC only) | 38 | 20.00 (52.63) |
| PHI <5% | 38 | 17.00 (44.74) |
| PHI <1% | 38 | 1.00 (2.63) |
| Water sites in village | 38 |  |
| None |  | 6.00 (15.79) |
| Beach |  | 14.00 (36.84) |
| Landing site only |  | 18.00 (47.37) |
| Public latrine in village | 38 | 24.00 (63.16) |

**Figure S1.**
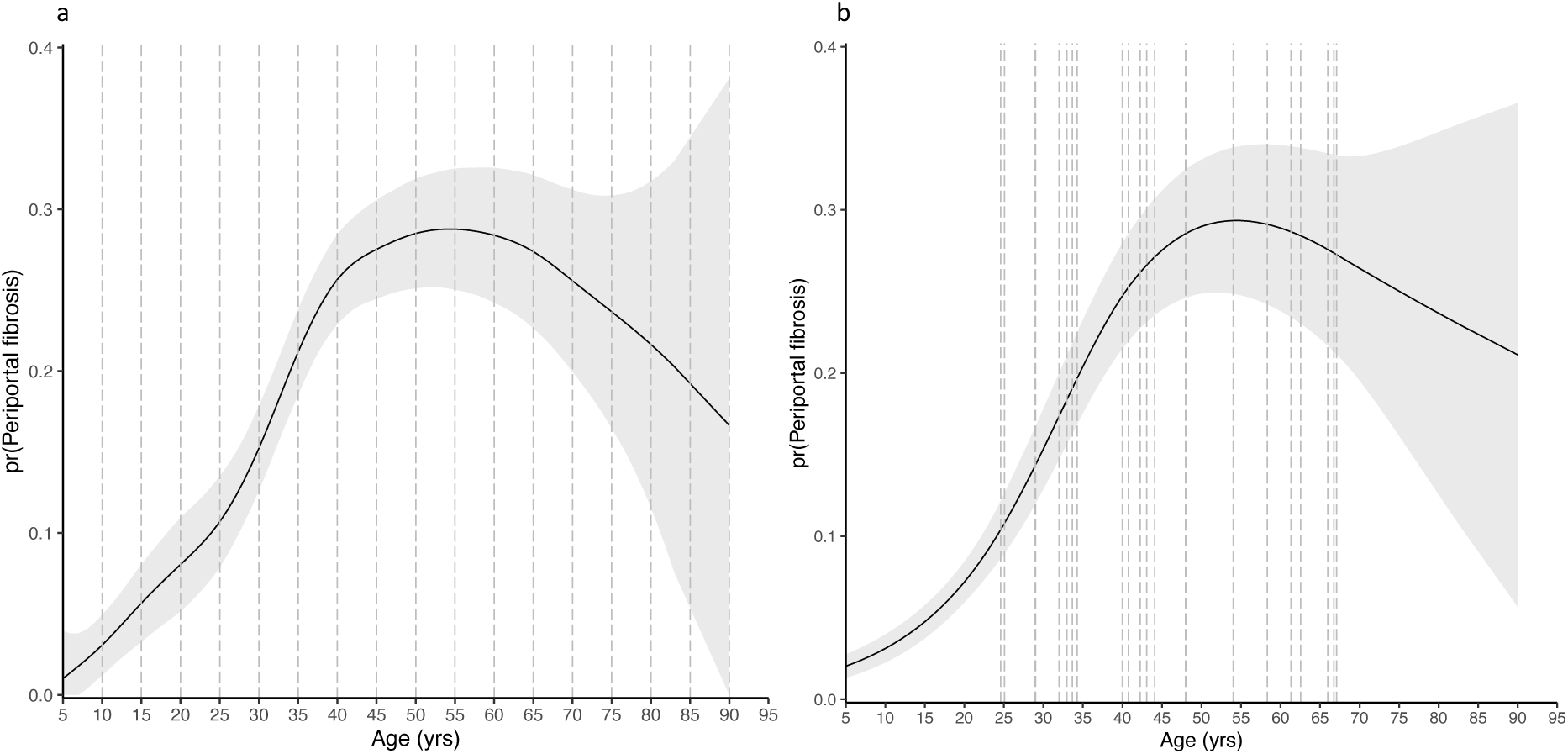
Generalised Additive Model (GAM) with fitted value (logit) of PPF over age. **a**. A GAM is shown with knots placed at every 5 years on the x-axis (age). **b**. A GAM is shown with 22 knots using the “freeknotsplines package in R’. It uses a random search algorithm and generalised cross validation (GCV) for selection of optimised knots numbers and their placement.^7^

**Figure S2.**
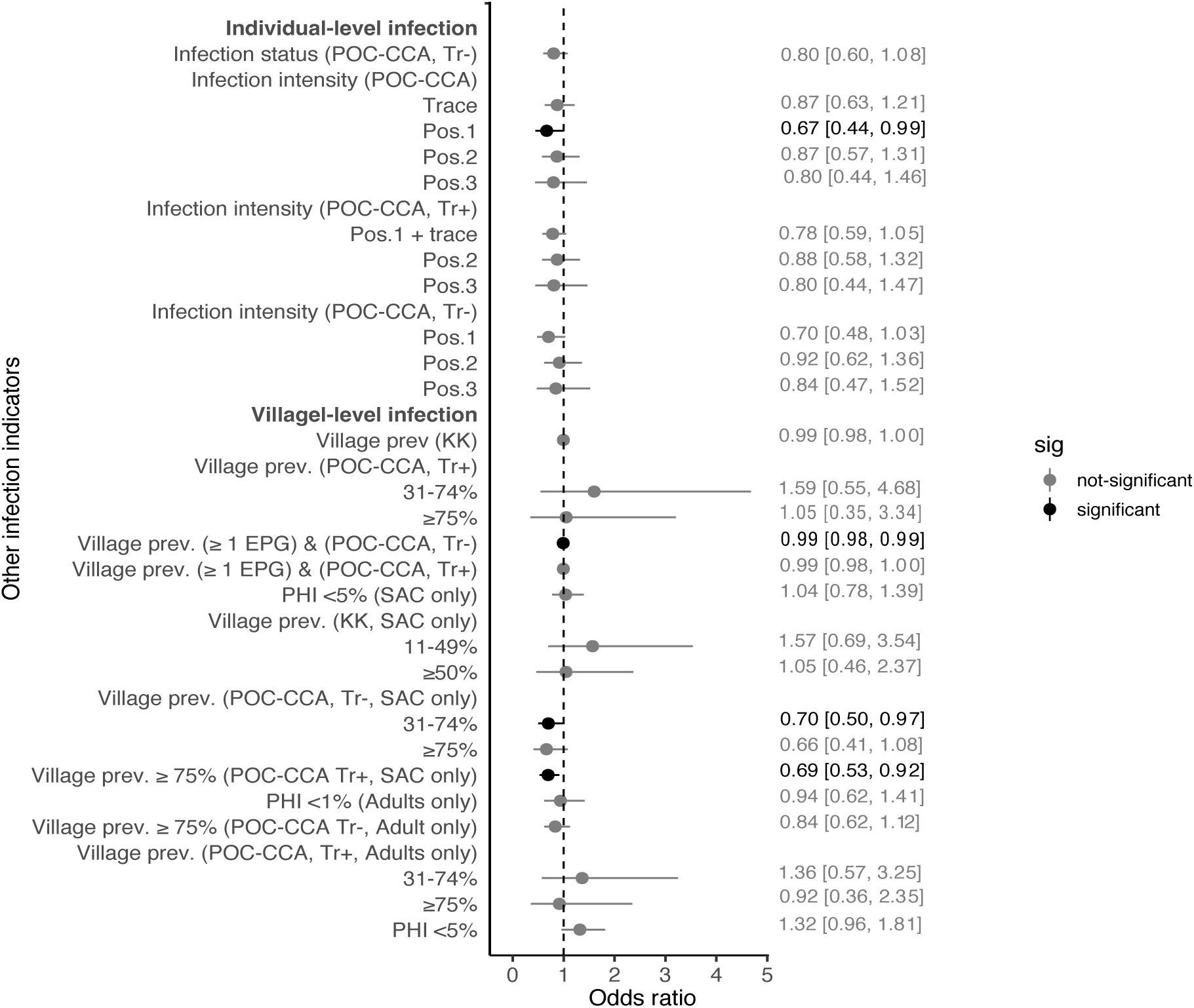
Fully adjusted models with alternative infection indicators. Alternative infection indicators, each representing a separate fully adjusted model, are shown. For each adjusted model, infection status (POC-CCA, Tr+) in Figure 6 was replaced by the other individual-level infection indicators each time while PHI <5% (Adults only) in Figure 6 was replaced by village-level infection indicators at the same time. All models were adjusted for age, gender, religion, tribe, all the medical history indicators, ultrasound-detected chronic hepatitis/early cirrhosis-like disease, malaria diagnosis, water contact, alcohol consumption and smoking status, occupation and all the household level variables in Figure 5. VIFs were between 1·1-2·3. The mean area under the ROC (10-fold CV) in each adjusted model ranged from 0·80-0·83.

**Figure S3.**
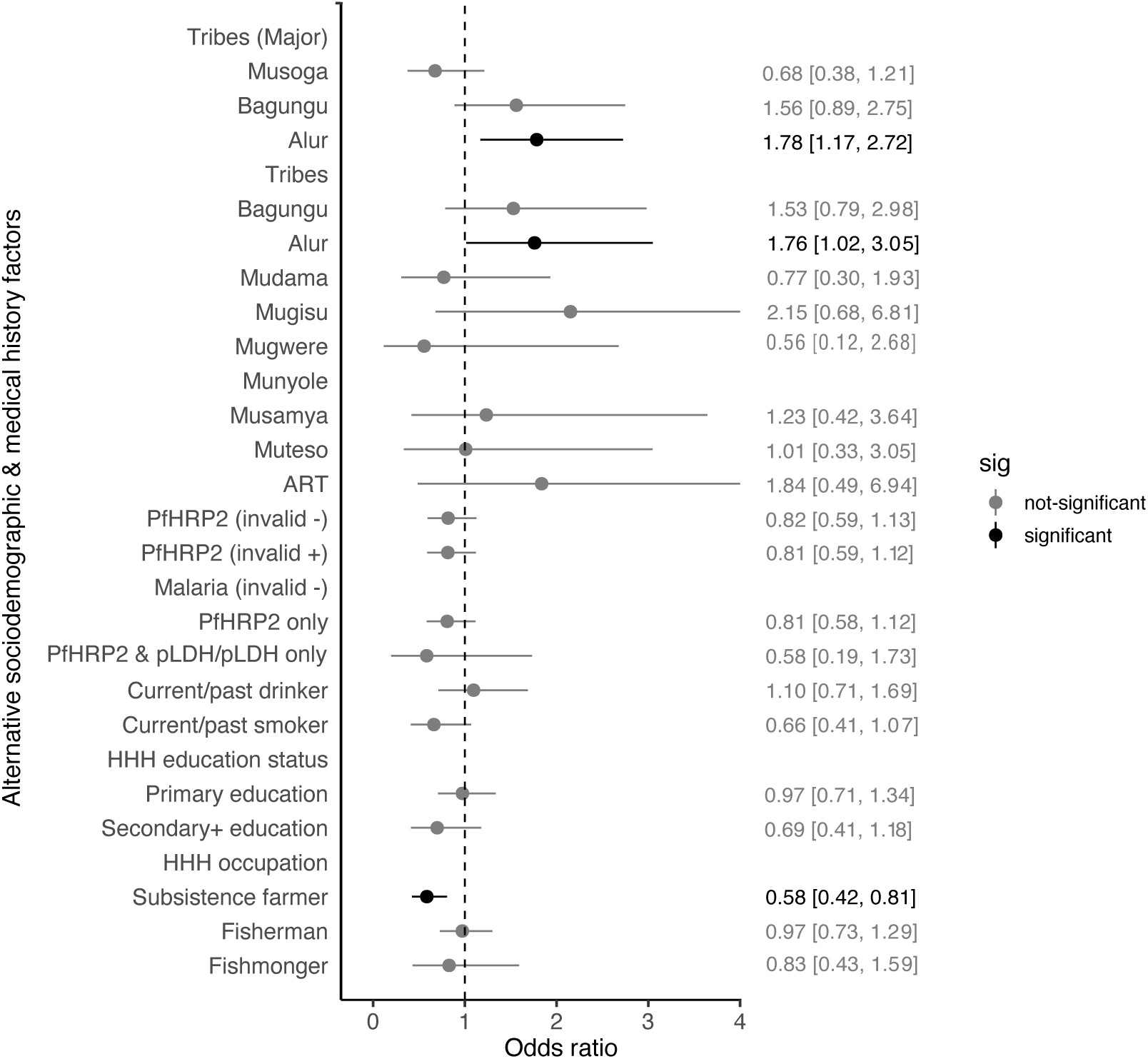
Fully adjusted models with alternative comorbidities and sociodemographic factors. Alternative definitions of sociodemographic and morbidity indicators, especially malaria, are shown where each represent a separate fully adjusted model. Each model was adjusted for age, gender, religion, tribe, all the medical history indicators, ultrasound-detected chronic hepatitis/early cirrhosis-like disease, malaria diagnosis, water contact, current drinker and current smokers, occupation and all the household level variables in Figure 5. VIFs were between 1·1-4·6. The mean area under the ROC (10-fold CV) in each adjusted model ranged from 0·81-0·84. PfHRP2 = Plasmodium falciparum antigen histidine rich protein 2. pLDH = parasite lactase dehydrogenase (generic to all malaria species). For pLDH only, there were only four cases.

**Figure S4.**
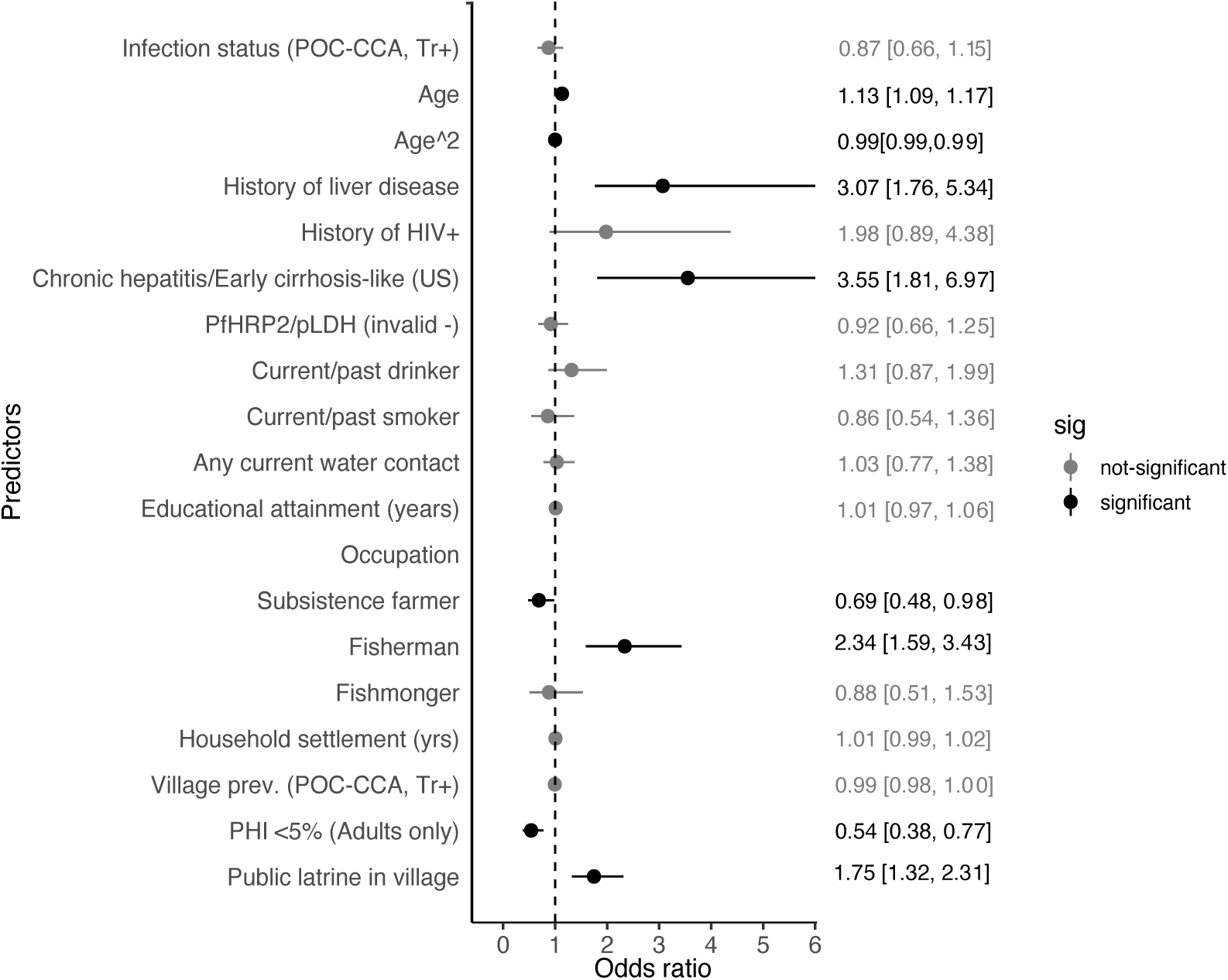
Subgroup analysis of participants from Pakwach District. Obs.= 925. Black represents significant relationships of p-value ≤0·05. VIFs were <10. The mean area under the ROC (10-fold CV) was 0·81. PfHRP2 = Plasmodium falciparum antigen histidine rich protein 2. pLDH = parasite lactase dehydrogenase (generic to all malaria species). For pLDH only, there were only four cases. US = Ultrasound.

**Figure S5.**
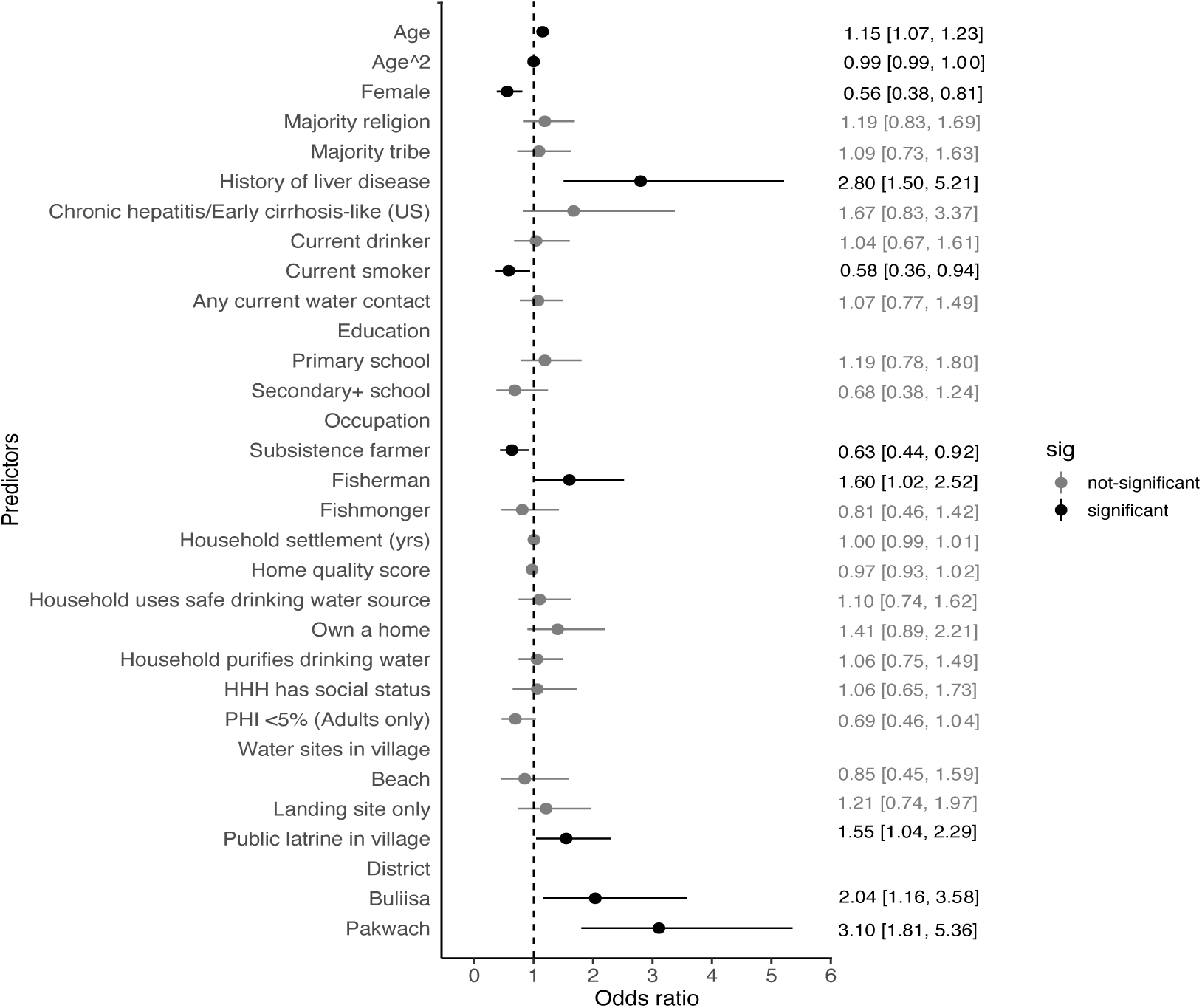
Subgroup analysis of adult participants. Obs. 1426. Black represents significant relationships of p- value≤0·05. VIFs were <10. The mean area under the ROC (10-fold CV) was 0·74. US = Ultrasound

**Figure S6.**
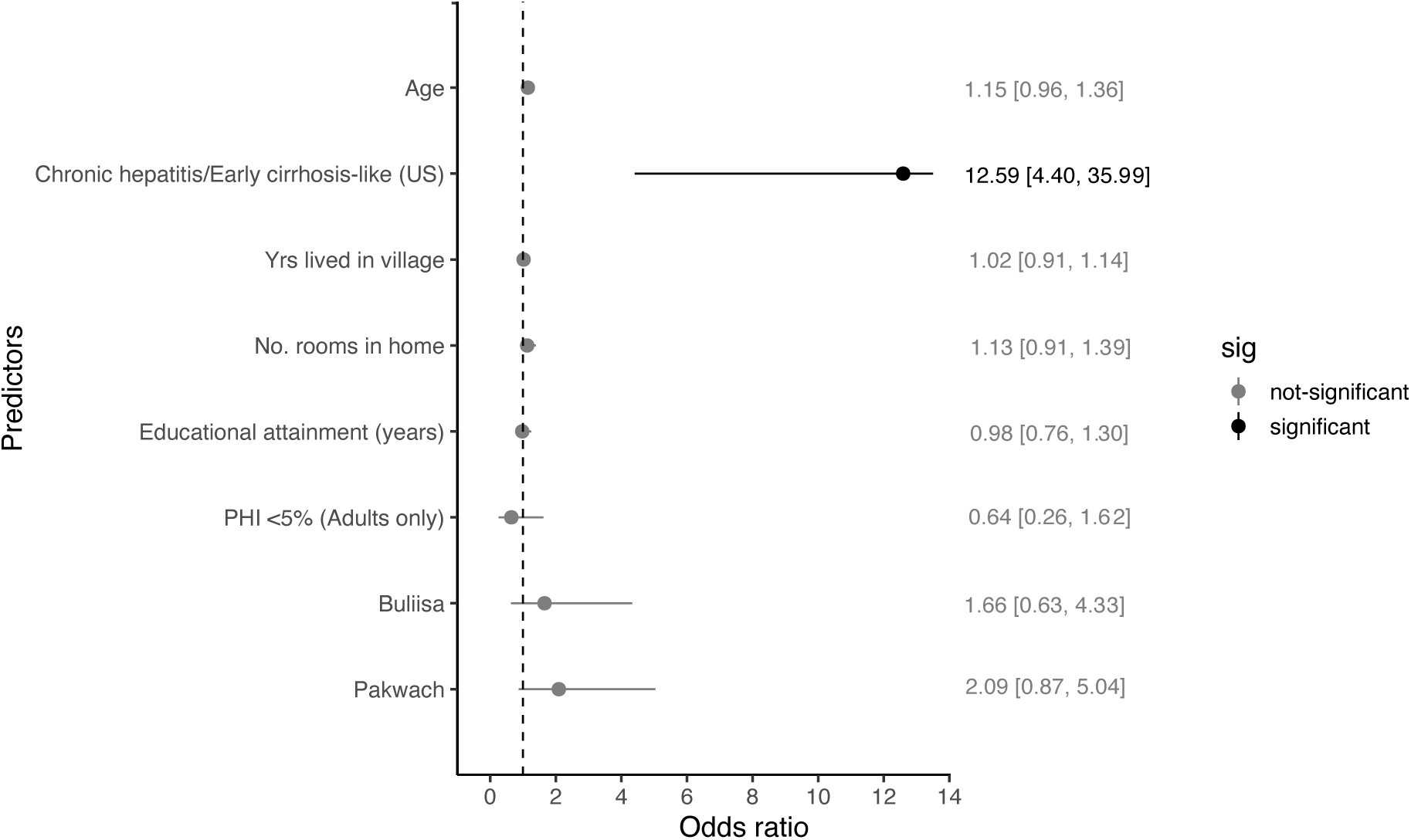
Subgroup analysis of children participants. Obs=1408. Black represents significant relationships of p-value≤0·05. VIFs were l<10. The mean area under the ROC (10-fold CV) was 0·70. US = Ultrasound

**Figure S7.**
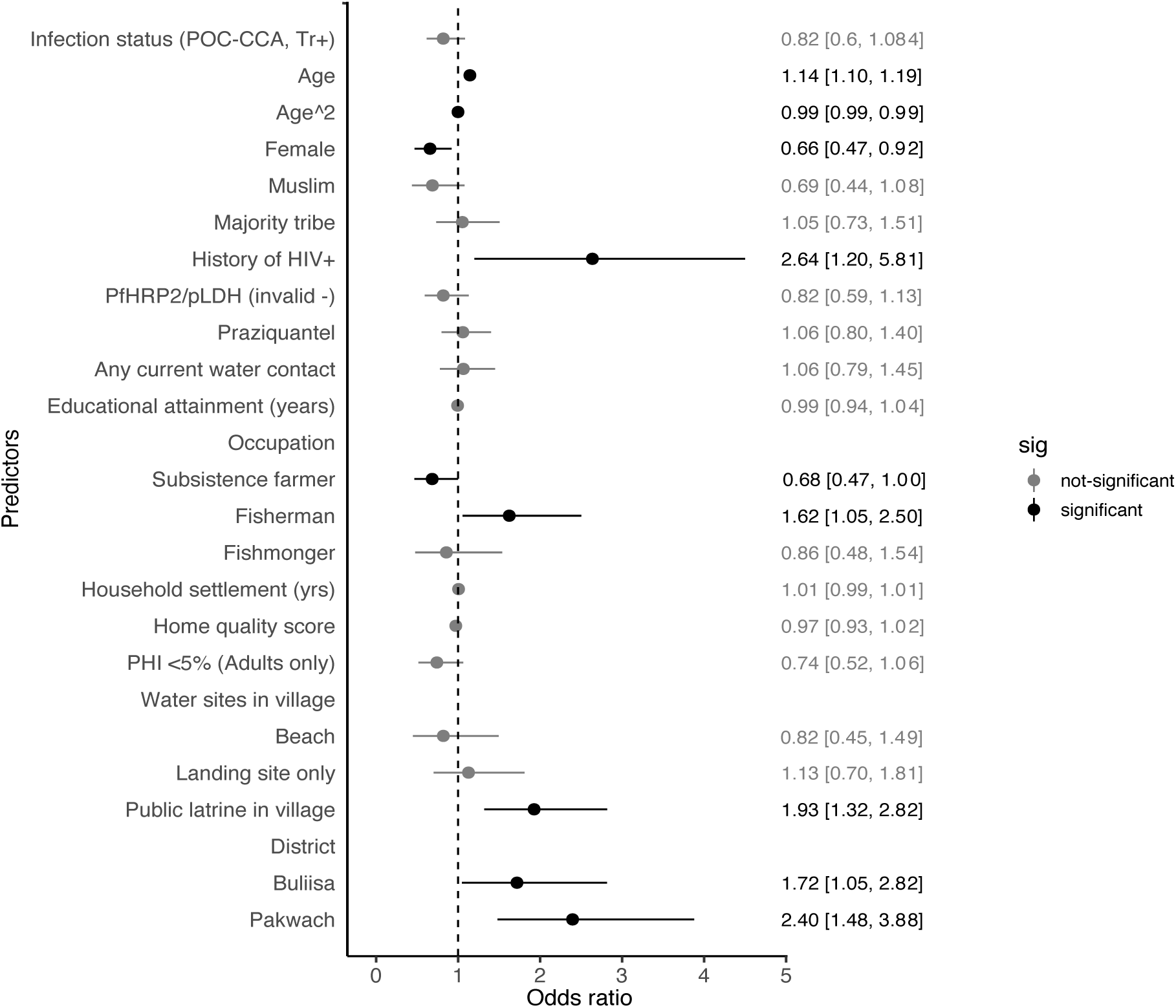
Sensitivity analysis of XY liver patterns. Obs.=2834. Study participants with liver pattern XY (the presence of a comorbidity including ultrasound-detected chronic hepatitis/early cirrhosis-like disease, fatty-like liver or a history of any liver diseases were recoded as zero (no PPF). Black represents significant relationships of p-value≤0·05. VIFs were <10. The mean area under the ROC (10-fold CV) was 0·81. PfHRP2 = *Plasmodium falciparum* antigen histidine rich protein 2. pLDH = parasite lactase dehydrogenase (generic to all malaria species). For pLDH only, there were only four cases.

